# Strain- and vaccine-specific effects of serum antibodies in the protection of intestinal SARS-CoV-2 infection

**DOI:** 10.1101/2025.03.24.25324570

**Authors:** M.D. Cherne, D. Snyder, B. Sidar, K. Blackwell, B. Jenkins, S. Huang, T.A. Sebrell, J.F. Hedges, J.R Spence, C.B Chang, J.N. Wilking, S.T. Walk, M.A. Jutila, E.K. Loveday, D. Bimczok

## Abstract

**Background:** Severe acute respiratory syndrome coronavirus 2 (SARS-CoV-2) infection remains a public health challenge worldwide. The gastrointestinal tract has emerged as an important site of infection and has been implicated as a reservoir for long-term infection, particularly for post-acute COVID-19 syndrome. However, whether vaccine-induced systemic antibodies can prevent intestinal infection with SARS-CoV-2 is unclear. Compared to Vero cells commonly used to assess SARS-CoV-2 neutralization, the intestinal epithelium has a functional interferon response and expresses higher levels of ACE2, enzymes, and antibody-binding Fc receptors that may impact SARS-CoV-2 immune elimination.

**Methods:** We evaluated the potential of antibodies from both naturally infected and vaccinated human subjects to inhibit SARS-CoV-2 infection of the intestinal epithelium. Serum samples were collected from human volunteers who had undergone natural infection with SARS-CoV-2 in 2020 (n=5) or who had received the Pfizer BNT162b2 COVID-19 vaccine (n=13). Banked sera collected in 2016 served as negative controls (n=2). SARS-CoV-2 (WA01, Delta or Omicron) was pre-treated with sera and then used to infect iPSC-derived human intestinal organoids (HIO) or Caco-2 colonic epithelial cells, and SARS-CoV-2 infection was quantified by plaque assay, PCR, or immunofluorescence (IF) after 48-96 h.

**Results:** Both HIOs and Caco-2 cells supported robust infection with SARS-CoV-2. In HIOs, pretreatment of SARS-CoV-2 with a high titer post-vaccine serum completely blocked replication of WA01. Similarly, sera from both naturally infected donors collected in 2020 and sera from individuals who had received a BNT162b2 vaccine significantly inhibited replication of the WA01 strain in Caco-2 cells. In contrast, none of the sera significantly inhibited infection with the Delta variant of SARS-CoV-2. For Omicron, only sera from individuals who had received an Omicron-based vaccine significantly inhibited infection with SARS-CoV-2 in the plaque assay. Across all virus types, sera from individuals who had received Omicron-based BNT162b2 boosters were the most effective at reducing infection in Caco-2 cells.

**Conclusion:** Our results suggest that vaccine-induced antibody responses to SARS-CoV-2 are protective in the gut. Our study also supports previous reports indicating that SARS-CoV-2 vaccines need to be adapted to circulating virus strains to convey full protection from infection.

## Introduction

The COVID-19 pandemic caused by infection with the severe acute respiratory syndrome coronavirus 2 (SARS-CoV-2) has caused more than 750 million infections and more than seven million deaths worldwide between 2020 and 2024 [1, 2], and SARS-CoV-2 remains a significant cause of respiratory illness today. In addition to the respiratory tract, the gastrointestinal tract is considered an important site of infection [3–6] and has been implicated as a reservoir for long-term viral persistence, particularly in the context of post-acute COVID-19 syndrome [7, 8]. Multiple studies have demonstrated that enterocytes in the small intestine and colon can harbor SARS-CoV-2, even after respiratory symptoms have resolved [5, 9].

Infected enterocytes also may serve as a viral reservoir that stimulates memory B cells in the gut, contributing to the evolution of the immune response to SARS-CoV-2 [10]. Enterocytes are among the cell types that express the highest level of the SARS-CoV-2 receptor ACE2 in the human body [11]. After engagement of ACE2, SARS-CoV-2 can enter cells following spike protein activation on the cell surface by the transmembrane protease, serine 2 (TMPRSS2), or in endosomal compartments via cathepsin L, and intestinal epithelial cells express both proteases [12, 13]. Therefore, intestinal epithelial cells are highly susceptible to SARS-CoV-2.

Antibody-mediated protection is considered a key mechanism of defense against SARS-CoV-2 [14], and vaccine induced responses are typically stronger than those triggered by natural infection [14, 15]. Therefore, multiple commercial vaccines were developed that induce antibody responses to SARS-CoV-2 [16], mostly targeting the SARS-CoV-2 spike (S) protein and thus leading to inhibition of viral ACE-2 binding [12]. One of the most widely distributed vaccines for SARS-CoV-2 is BNT162b2, a nanoparticle-based mRNA vaccine produced by BioNTech and Pfizer that encodes the S glycoprotein [14, 17, 18]. BNT162b2 demonstrated an excellent safety profile and strong protection from natural infection in clinical trials and has been updated multiple times to induce improved immunity to new viral variants of concern, i.e., Omicron strains [19]. Importantly, the BTN162b2, which is applied intramuscularly, induces both systemic and mucosal antibody responses [20], However, whether vaccine-induced antibodies prevent intestinal infection with SARS-CoV-2 has not been systematically investigated.

Notably, anti-SARS-CoV-2 antibody responses are most commonly measured either by ELISA or based on virus neutralization assays performed in VeroE6 cells, which are derived from monkey kidneys [21]. VeroE6 cells lack type I interferon production [22], but express high levels of the SARS-CoV-2 receptor ACE2 [23, 24] and thus have an unphysiologically high susceptibility to SARS-CoV-2 infection. Therefore, data from Vero cells cannot simply be extrapolated to all other cell types without experimental verification. Human intestinal epithelial cells have intact type I interferon production, but unlike VeroE6 cells, they express antibody receptors (FcRs) including FcRn and pIgR, which may provide alternative pathways for viral binding and entry [25, 26].

In this study, we evaluated the potential of serum antibodies from both naturally infected and vaccinated human subjects to inhibit infection of the intestinal epithelium with different strains of SARS-CoV-2 using organoid and cell line models. We used both pluripotent stem-cell derived human intestinal organoids, which maintain the cellular diversity of the human intestinal epithelium [27], and Caco-2 cells, an immortalized human colonic cell line that is known to support robust SARS-CoV-2 replication [28–31]. We found that antisera from individuals that had been naturally infected with SARS-CoV-2 or had received the BNT162b2 COVID-19 vaccine blocked infection of intestinal organoids and Caco-2 cells with the WA-01 strain of SARS-CoV-2, but showed a variable efficacy for inhibition of intestinal infection with the SARS-CoV-2 Delta and Omicron strains. Sera from individuals immunized with Omicron-based COVID-19 vaccines showed the highest protection against all SARS-CoV-2 strains. Overall, our results highlight that antibody protection from intestinal SARS-CoV-2 infection depends on virus strain and vaccine antigens and may involve some cell-type specific effects.

## Materials and Methods

### Cells lines and maintenance

Caco-2 colonic epithelial cells were purchased from ATCC (#HTB-37; Manassas, VA, USA) and were maintained in DMEM supplemented with 20% FBS, penicillin, streptomycin, L-glutamine, and fungizone, with passaging every 4-6 days. VeroE6 cells also were obtained from ATCC (#CRL-1586) and grown in DMEM (ThermoFisher, Waltham, MA, USA) supplemented with 10% FBS (Atlanta Biologicals, Minneapolis, MN, USA), penicillin, and streptomycin (ThermoFisher, Waltham, MA, USA).

### Human intestinal organoids

Human Intestinal Organoids (HIOs), prepared as described previously from H9 human embryonic stem cells [27], were cultured in Dr. Jason Spence’s laboratory at the University of Michigan. Early-stage HIOs were mailed to Montana State University with over-night shipping and were immediately plated into 24-well plates at a density of ∼50 HIOs in 50 µL Matrigel patties. The wells containing HIOs were incubated at 37°C in intestinal growth media which consisted of a basal media (containing penicillin, streptomycin, Glutamax, HEPES, B27, and Advanced DMEM/F12) supplemented with noggin, R-spondin1, and epidermal growth factor (EGF). The intestinal growth media was changed every other day. The HIOs were passaged once a week to prevent collapse from mesenchyme overgrowth.

### Human sera

Serum samples were collected with written and witnessed informed consent from adult volunteers with approval from the Institutional Review Board at Montana State University (protocol # JH041020), as previously described [15]. Participants for the study were recruited between April 10^th^, 2020, and November 30th, 2023, by way of informative flyers posted in two buildings at Montana State University, and by word of mouth. These included (1) five individuals with previous positive SARS-CoV-2 tests due to natural infection in 2020/21, before vaccines were available (“natural”), (2) six individuals who had received the monovalent COVID-19 mRNA vaccine Comirnaty/BNT162b2 that was based on the Wuhan-Hu-01 strain from Pfizer in 2021 (“original” [17]), and (3) six individuals who had either received an updated bivalent mRNA vaccine based on Wuhan-Hu-1 isolate and the Omicron variants BA.4 and BA.5 or a vaccine based on the Omicron XBB 1.5 lineage from Pfizer in 2023 (“Omicron”). Details on the collected sera are provided in **Table 1**. The sera from the natural infection group and the original Pfizer vaccine group were analyzed and documented in a previous study by our team [15]. Informed consent was obtained from all participants. Deidentified, archived serum samples from two human subjects obtained in 2016, well before the start of the COVID-19 pandemic, were accessed between May 15, 2022, and June 30^th^, 2024, and used as negative controls (“negative”). One tube of blood was collected in a BD Vacutainer SST Venous Blood Collection Tube from each patient per sampling day. Tubes were centrifuged at 1,150 *g* for 10 min to separate the serum. Aliquots of sera were delivered directly to our BSL-3 facility and stored at 4° C until use in experiments.

**Table 1:**
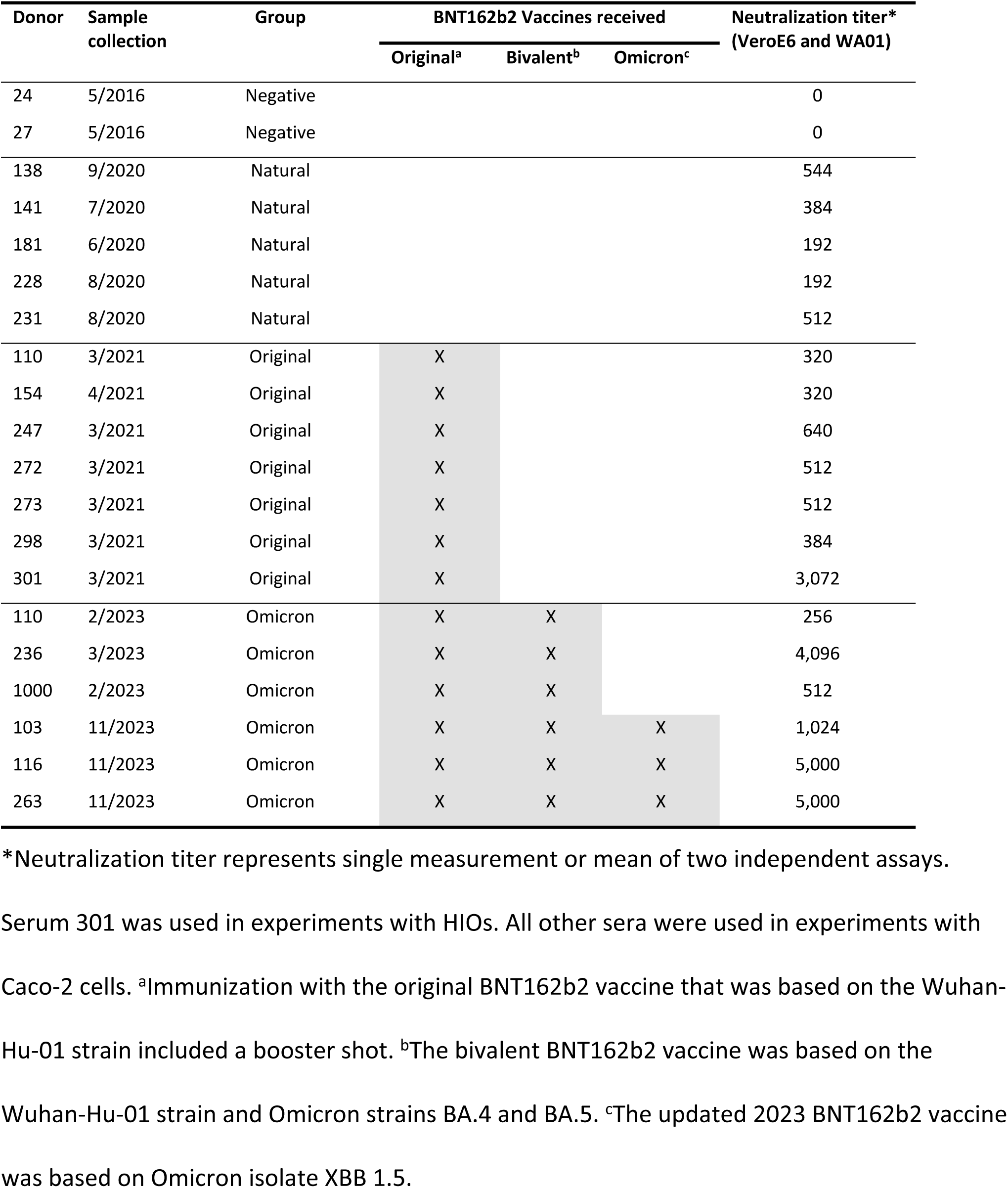
Serum donors and neutralization titers.

| Donor | Sample collection | Group | BNT162b2 Vaccines received |  |  | Neutralization titer*<br>(VeroE6 and WA01) |
| --- | --- | --- | --- | --- | --- | --- |
|  |  |  | Original<sup>a</sup> | Bivalent<sup>b</sup> | Omicron<sup>c</sup> |  |
| 24 | 5/2016 | Negative |  |  |  | 0 |
| 27 | 5/2016 | Negative |  |  |  | 0 |
| 138 | 9/2020 | Natural |  |  |  | 544 |
| 141 | 7/2020 | Natural |  |  |  | 384 |
| 181 | 6/2020 | Natural |  |  |  | 192 |
| 228 | 8/2020 | Natural |  |  |  | 192 |
| 231 | 8/2020 | Natural |  |  |  | 512 |
| 110 | 3/2021 | Original | X |  |  | 320 |
| 154 | 4/2021 | Original | X |  |  | 320 |
| 247 | 3/2021 | Original | X |  |  | 640 |
| 272 | 3/2021 | Original | X |  |  | 512 |
| 273 | 3/2021 | Original | X |  |  | 512 |
| 298 | 3/2021 | Original | X |  |  | 384 |
| 301 | 3/2021 | Original | X |  |  | 3,072 |
| 110 | 2/2023 | Omicron | X | X |  | 256 |
| 236 | 3/2023 | Omicron | X | X |  | 4,096 |
| 1000 | 2/2023 | Omicron | X | X |  | 512 |
| 103 | 11/2023 | Omicron | X | X | X | 1,024 |
| 116 | 11/2023 | Omicron | X | X | X | 5,000 |
| 263 | 11/2023 | Omicron | X | X | X | 5,000 |

\*Neutralization titer represents single measurement or mean of two independent assays.
Serum 301 was used in experiments with HIOs. All other sera were used in experiments with Caco-2 cells. <sup>a</sup>Immunization with the original BNT162b2 vaccine that was based on the Wuhan-Hu-01 strain included a booster shot. <sup>b</sup>The bivalent BNT162b2 vaccine was based on the Wuhan-Hu-01 strain and Omicron strains BA.4 and BA.5. <sup>c</sup>The updated 2023 BNT162b2 vaccine was based on Omicron isolate XBB 1.5.

### SARS-CoV-2 strains and production

The following strains of SARS-CoV-2 were obtained from BEI Resources (Manassas, VA, USA): WA01 (NR-52281), Delta AY.1 (NR-55691) or Omicron BA.1 (NR-56461). Viral stocks were prepared on E6 Vero cells in DMEM supplemented with 2% FBS and 1% penicillin/streptomycin. Media supernatants from infected cell cultures showing extensive cytopathic effect were collected and centrifuged at 1,000 *g* for 5 min to remove cellular debris. The clarified viral supernatant was titered by plaque assay and then used for all infection experiments.

### Determination of SARS-CoV-2 neutralization titers of human sera in VeroE6 cells

The neutralization titers of human sera were determined based on infection of VeroE6 cells with SARS-CoV-2 strain WA01 in BSL-3 containment, as previously described [15, 32]. Vero E6 cells were seeded at 4 x10^4^ cells per well in 96-well flat-bottom plates 24 h before the experiment to achieve confluent monolayers. The sera were serially diluted from 1:16 to 1:2,048 and mixed with 1000 TCID50 of SARS-CoV-2 (WA strain, BEI Resources) in DMEM. The mixture was incubated for 1 h then applied to the VeroE6 cell monolayers. The plates were incubated for 48 h, then the supernatant fluids were removed, and the cell monolayers were stained with (0.5%) methylene blue in 70% ethanol. The neutralization titers were determined as the inverse of the serum dilution that provided 50% protection of the monolayer, as previously described [15].

### Serum inhibition of SARS-CoV-2 infection in human intestinal organoids (HIOs)

HIOs were prepared for SARS-CoV-2 infection by first replating selected stocks into a 48-well plate so that each well contained approximately the same heterogeneity and number of organoids (approximately 5-10). 48 h before the infection, the intestinal growth media was replaced with differentiation media in which the noggin and R-spondin are replaced by 1% FBS. To prepare the HIOs for infection, a Matrigel patty containing the HIOs was relocated to a petri dish containing basal media under the dissecting scope with a p1000 tip cut to a wider bore with a sterile razorblade. The HIOs were gently removed from the Matrigel patty with scalpel and needle and the excess Matrigel and mesenchyme was trimmed. The HIOs were then bisected with a sterile scalpel and transferred into screwcap tubes with a p200 pipettor and resuspended in PBS. Cells were counted in a representative organoid and all organoids were transferred to the BSL3 for infection. Human serum samples were diluted to 1:128 in DMEM and mixed 1:2 for a final serum concentration of 1:256 with WA01 (NR-52281) to achieve a final MOI of 10. This mixture was transferred to each tube of HIOs and incubated for 90 min at 37 °C. Organoids were washed by adding 500 µL DMEM and then centrifuging at 200 *g* for 3 min.

Liquid was carefully removed by pipette and 40-50 µl of Matrigel was added to each tube of HIOs, on ice. Matrigel-HIO mixtures were then transferred to 48-well plates, Matrigel was allowed to solidify at RT for 10 min, and then 300 µl of differentiation media was added on top of the Matrigel. HIOs were incubated at 37 °C for 0, 48 or 72 h. Supernatants were collected for plaque assay or RNA extraction and HIOs were lysed in RLT buffer for RNA extraction.

### Serum inhibition of SARS-CoV-2 infection in Caco-2 cells

Confluent monolayers of Caco-2 cells were grown on glass-bottom 96-well plates. Before inoculation, three representative wells of Caco-2 cells were trypsinized and counted to obtain an average cell number per well for determination of MOI. In BSL3 containment, human sera were diluted to a final concentration of 1:256 by mixing a 1:128 dilution of serum with an equal volume of neat or 1:10 dilutions of WA01, Delta AY.1, or Omicron BA.1, respectively. Serum-treated virus was pre-incubated for 1 h before adding the virus-serum mixture to the Caco-2 cells at an MOI of 1. Infected Caco-2 cells were cultured for 24, 28, 72 or 96 h depending on the experiment. Supernatants were collected for plaque assays and Caco-2 monolayers were fixed for immunofluorescence analysis.

### Quantification of viral titers by plaque assay

SARS-CoV-2 titers were determined using plaque assays with VeroE6 cells, as previously described [33]. Briefly, Vero E6 cells were cultured in DMEM with 10% FBS, penicillin, streptomycin and plasmocin. For plaque assays, VeroE6 cells were seeded onto 6-well plates and grown to confluence overnight and then were incubated with serial diluted viral inoculum for 1 h. Following inoculation, cells were over-layered with 1% methylcellulose (Sigma-Aldrich, St. Louis, MO, USA), DMEM supplemented with 2% FBS and 1% penicillin/streptomycin and incubated for 3–4 days. Cells were then fixed and stained with 0.5% methylene blue (Sigma-Aldrich, St. Louis, MO, USA) in 70% ethanol. Plaques were counted and the titer was calculated based on the volume of inoculum plated and dilution counted.

### Quantification of SARS-CoV-2 infection by quantitative RT-PCR

To quantify SARS-CoV-2 in culture supernatants or organoids, viral RNA was extracted from culture supernatants using the QIA®Amp Viral RNA Mini kit (Qiagen) following the manufacturer’s instructions. Viral genomes were then quantified in a single-step RT-PCR reaction using primers for SARS-CoV-2 envelope (E) gene: (forward: ACAGGTACGTTAATAGTTAATAGCGT; reverse: ATATTGCAGCAGTACGCACACA) and a TaqMan probe (FAM-ACACTAGCCATCCTTACTGCGCTTCG-BHQ1), as previously described [33, 34], and the Quanta Bio ToughMix Master Mix. For the detection of subgenomic SARS-CoV-2 RNA in cell pellets, the forward primer was replaced by an alternative primer for the leader sequence, originally designed by Wölfel et al. [35] (forward-subG: CGATCTCTTGTAGATCTGTTCTC). GAPDH was used as a house keeping gene to normalize cell-associated viral gene expression, which was analyzed using the Delta Delta cT method. Gene expression of GAPDH was determined using a two-step reaction with SYBR green for quantification, following our published protocol [36]. PCR reactions were run on a QuantStudio real time PCR system.

### Quantification of SARS-CoV-2 infection by immunofluorescence staining

To analyze the effect of serum neutralization on SARS-CoV-2 infection of Caco-2 cells using immunofluorescence staining, plates with Caco-2 cells from the SARS-CoV2 infection and serum neutralization experiments were fixed with 4% paraformaldehyde. Cells were then incubated with blocking buffer (DPBS with 10% FBS, 0.2% Triton X-100, 0.1% BSA, and 0.05% Tween) for 1 h at room temperature on a shaking platform. A primary antibody for the SARS-CoV-2 nuclear protein (NP, Invitrogen MA1-7403) was added at a 1: 100 dilution in blocking buffer and incubated over night at 4 °C. After three washing steps with PBS supplemented with 0.05% Tween 20, cells were incubated with a secondary goat anti-mouse IgG2b-Alexa 555 antibody (Southern Biotechnologies, Birmingham, AL) for 45 min at room temperature, followed by a 5 min incubation step with (DAPI, 5 µM). The plates were then washed again with PBS.

Whole well scans were performed on a Leica Stellaris 8 Confocal Scanning Laser Microscope (CSLM) with HC PL APO CS2 10x/0.40 DRY objective and a Leica DMI8 widefield microscope HC PL FLUOTAR 10x/0.32 DRY objective and K8 monochrome camera using LAS X software versions 4.4.0 and 4.5.0 (Leica Microsystems Inc. Wetzlar, Germany). Fluorophores were excited with a diode 405 nm laser (DAPI emission: 424–535nm), a white light laser at 553 nm (Alexa555 emission: 558–737nm) for CSLM, and multiline tunable LED light source and a quadband filter cube with 390/440 nm (DAPI), 555/590 nm (Alexa555) excitation/emission settings for widefield microscope. For image analysis, stained cells were analyzed with Imaris® v10.0 by surface analysis, followed by automated identification and counting of DAPI or Alexa555 positive florescent objects (**Supplemental Fig. 1**). Cell nuclei for total cell count were defined as DAPI fluorescent objects, with a single object predicted at 8.52 µm. Infected cells were defined as red fluorescent objects with a maximum diameter of 40 µm, and object splitting was performed above this threshold. Red fluorescent objects with no distinguishable overlapping DAPI signal, or that were over 60 µm in diameter were excluded.

### Statistical analyses

Statistical analyses were performed with GraphPad Prism® version 10.1.2. (GraphPad Software, Boston, MA). Data are shown as individual values, mean, and standard deviation (SD), and were tested for statistically significant differences using one-way ANOVA with Dunnet’s multiple comparison test or two-way ANOVA with Tukey’s multiple comparisons test. Simple non-linear regression was used to compare neutralizing activity in sera in VeroE6 and Caco-2 cells.

## Results

### Vaccine-induced antibodies protect human intestinal organoids from SARS-CoV-2 infection

Previous studies have demonstrated that SARS-CoV-2 can infect primary human enterocytes in multiple different organoid models [3, 29, 37–39]. In our hands, human intestinal organoids (HIOs) derived from H9 human embryonic stem cells (**Fig. 1A**) supported robust viral infection, with a >1 log-fold increase in SARS-CoV-2 plaque forming units (PFUs) at 72 h post infection with the WA01 strain (**Fig. 1B**).

**Figure 1:**
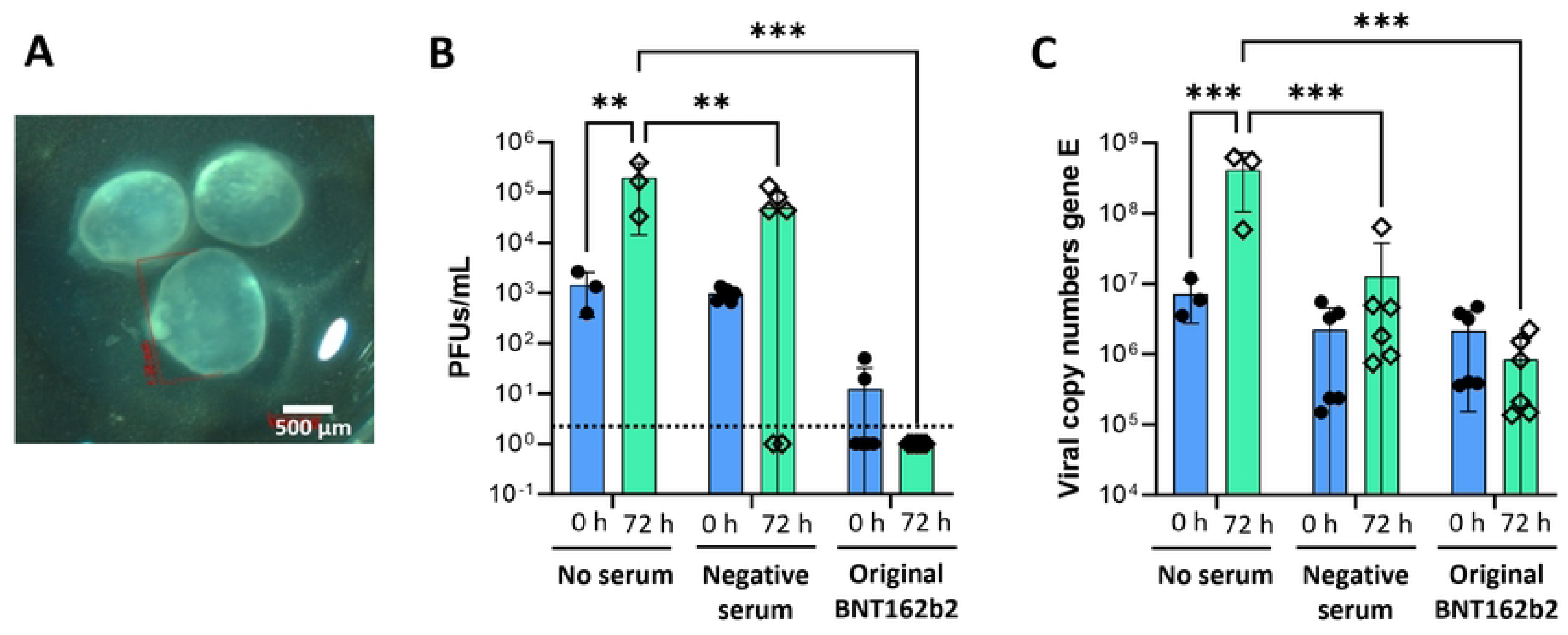
Serum inhibition of intestinal epithelial cell infection with SARS-CoV-2 in human intestinal organoids. (**A**) Phase contrast image of representative embryonic stem cell-derived human intestinal organoids (HIOs) prior to SARS-CoV-2 infection. (**B, C**) SARS-CoV-2 was mixed with sera diluted 1: 256 and then added at an MOI of 10 to HIOs that were prepared for infection by cutting them with a scalpel. HIOs were re-embedded in Matrigel after 1.5 h and then were analyzed at 72 h post infection. Pooled data from two experiments with three technical replicates each are shown, individual data points, mean ± SD. “No serum” samples were analyzed only in one experiment. 2-way ANOVA with Tukey’s multiple comparisons test. (**B**) Plaque assays of HIO culture supernatants were performed in VeroE6 cells. Dotted line indicates limit of detection. (**C**) SARS-CoV-2 in culture supernatants was analyzed using qRT-PCR for gene E. Copy numbers were calculated based on a standard curve.

To assess whether vaccine-induced antibodies could block infection, SARS-CoV-2 WA01 virus was pre-incubated with buffer, a control serum collected in 2016 prior to the COVID-19 pandemic, or a serum sample of a donor (#301) who had received the original Pfizer BNT162b2 COVID-19 vaccine in 2021 that showed a high serum neutralization titer in VeroE6 cells (1: 3,072; **Table 1**). The serum sample from the vaccinated donor, diluted 1: 256, significantly blocked SARS-CoV-2 replication in the HIOs, as evidenced by a reduction in PFUs in the organoid supernatant to a level that was below the limit of detection (*P* ≤ 0.001, **Fig. 1B**). In contrast, a negative control serum that had no neutralizing activity in VeroE6 cells allowed replication of SARS-CoV-2, as evidenced by an increase in PFUs over 72 h. However, viral replication was lower than for the untreated control (*P* ≤ 0.01; **Fig. 1B**). Quantitative PCR analysis for the SARS-CoV-2 E gene in culture supernatants showed similar results (**Fig. 1C**). Analysis of subgenomic RNA for the E gene in cell pellets confirmed productive SARS-CoV-2 replication in the HIOs as well as inhibition of SARS-CoV-2 replication by the serum from the vaccinated individual (**Supplemental Fig. 2**). These results support previous reports indicating that human intestinal epithelial cells are susceptible to SARS-CoV-2 replication, but that intestinal infection can be successfully blocked by neutralization with immune sera.

### SARS-CoV-2 WA01, Delta and Omicron strains replicate robustly in Caco-2 cells, but infect only a limited number of cells

To further investigate SARS-CoV-2 infection and antibody-mediated inhibition in the human intestine, we next explored Caco-2 cells as a high-throughput model of intestinal epithelial infection with SARS-CoV-2. Caco-2-cells are an epithelial line derived from a human colon carcinoma [40], and were shown to be susceptible to SARS-CoV-2 in previous studies by other groups [28–31]. Confirming these results, we show that Caco-2 cells were susceptible to infection with the SARS-CoV-2 WA01 strain and supported a >3 log-fold increase in PFUs over 96 h (**Fig. 2A**). Viral particle release increased from 0 h to 48 h and from 48 to 96 h for MOIs 0.1 and 1, but remained relatively stable between 48 h and 96 h for the MOI of 10. Viral titers peaked around 5 x10^6^ PFUs/mL after 96 h for all MOIs. Therefore, an MOI of 1 and a 48 h time point was chosen for all subsequent experiments.

**Figure 2:**
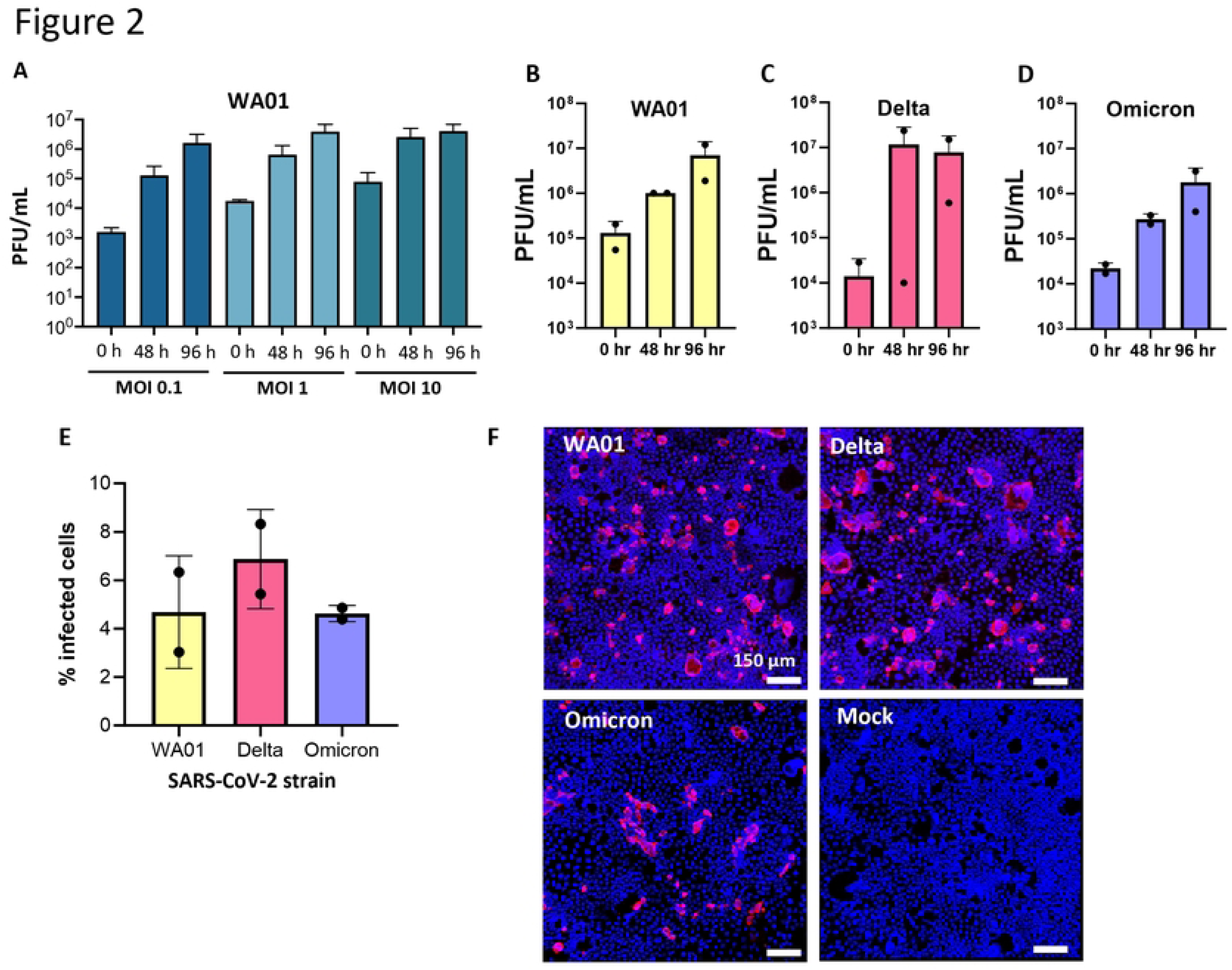
Robust replication of SARS-CoV-2 WA01, Delta and Omicron in Caco-2 intestinal epithelial cells. (**A**) Viral replication of SARS-CoV-2 WA01 after 48 and 96 h at different multiplicities of infection (MOIs). Mean ± of 2-3 wells. (**B-D**) Replication of SARS-CoV-2 WA01, Delta, and Omicron following inoculation with an MOI = 1 at 48 and 96 h. Data represent two independent experiments with two technical replicates each; mean ± SD. (**E**) Percentage of SARS-CoV-2-infected Caco-2 cells 48 h after inoculation of monolayers with different virus strains at an MOI of 1, determined by immunofluorescence analysis of viral nuclear protein and digital image analysis. Data represent two independent experiments with two technical replicates each (**F**) Representative immunofluorescence images of SARS-CoV-2-infected Caco-2 monolayers for viral nuclear protein (Alexa 555, red). DAPI nuclear stain is shown in blue. Bar = 150 µm.

Comparison of the SARS-CoV-2 strains WA01, Delta and Omicron showed that Caco-2 cells supported infection with all three strains, with slightly higher viral titers seen in the plaque assay with the Delta strain (**Fig. 2B-D**). Infection of the epithelial cells was confirmed using immunofluorescence staining of Caco-2 monolayers for the SARS-CoV-2 nuclear protein (NP). Interestingly, only a small proportion of the cells (<10%) showed evidence of viral infection with any of the strains (**Fig. 2E,F**). Again, the proportion of infected cells was generally higher following infection with SARS-CoV-2 Delta compared to the other strains (**Fig. 2E**). Caco-2 monolayers remained largely intact after SARS-CoV-2 infection. While no major cytopathic effects occurred within the time frame of the experiment (**Supplemental Fig. 3**), some syncytia formation was observed in the Caco-2 cells, consistent with previous reports [28].

### Collection of sera from individuals exposed to natural SARS-CoV-2 infection or different types of the Pfizer BTN162b2 vaccine

We next analyzed to what extent antibodies induced by natural SARS-CoV-2 infection and commercial COVID-19 vaccines can prevent infection of the intestinal epithelium. To that end, we collected sera from individuals with natural SARS-CoV-2 infection (n=5), sera from individuals who had received the original Pfizer BNT162b2 mRNA vaccine that was based on the Wuhan-Hu-01 strain in 2021 (n=6), and sera from individuals who had received updated vaccines containing Omicron sequences in early or late 2023 (n=6; **Table 1**). Sera collected from two individuals in 2016, prior to the COVID-19 pandemic, were used as negative controls. Notably, all serum samples except the negative control sera were shown to inhibit SARS-CoV-2 WA01 infection in routine virus neutralization assays in VeroE6 cells (**Table 1**). Prolonged storage of the sera for up to three years did not significantly affect antibody titers in the samples (**Supplemental Fig. 4A**). Overall, antibody titers tended to be highest after vaccination with the Omicron-based booster vaccines and lowest following natural infection, although these differences were not significant (**Supplemental Fig. 4B** and **Table 1**).

### Protective effects of sera from SARS-CoV-2 infection of intestinal epithelial cells depend on virus strain and vaccine type

We next compared serum inhibition of Caco-2 SARS-CoV-2 infection by the different types of sera for three strains of SARS-CoV-2: WA01, Delta AY.1 and Omicron BA.1. As expected, sera from both the original and Omicron-based BNT162b2 vaccinees significantly inhibited replication of the WA01 strain of SARS-CoV-2 in Caco-2 cells, with no PFUs detected with the Omicron-based vaccine serum (**Fig. 3A**).

**Figure 3:**
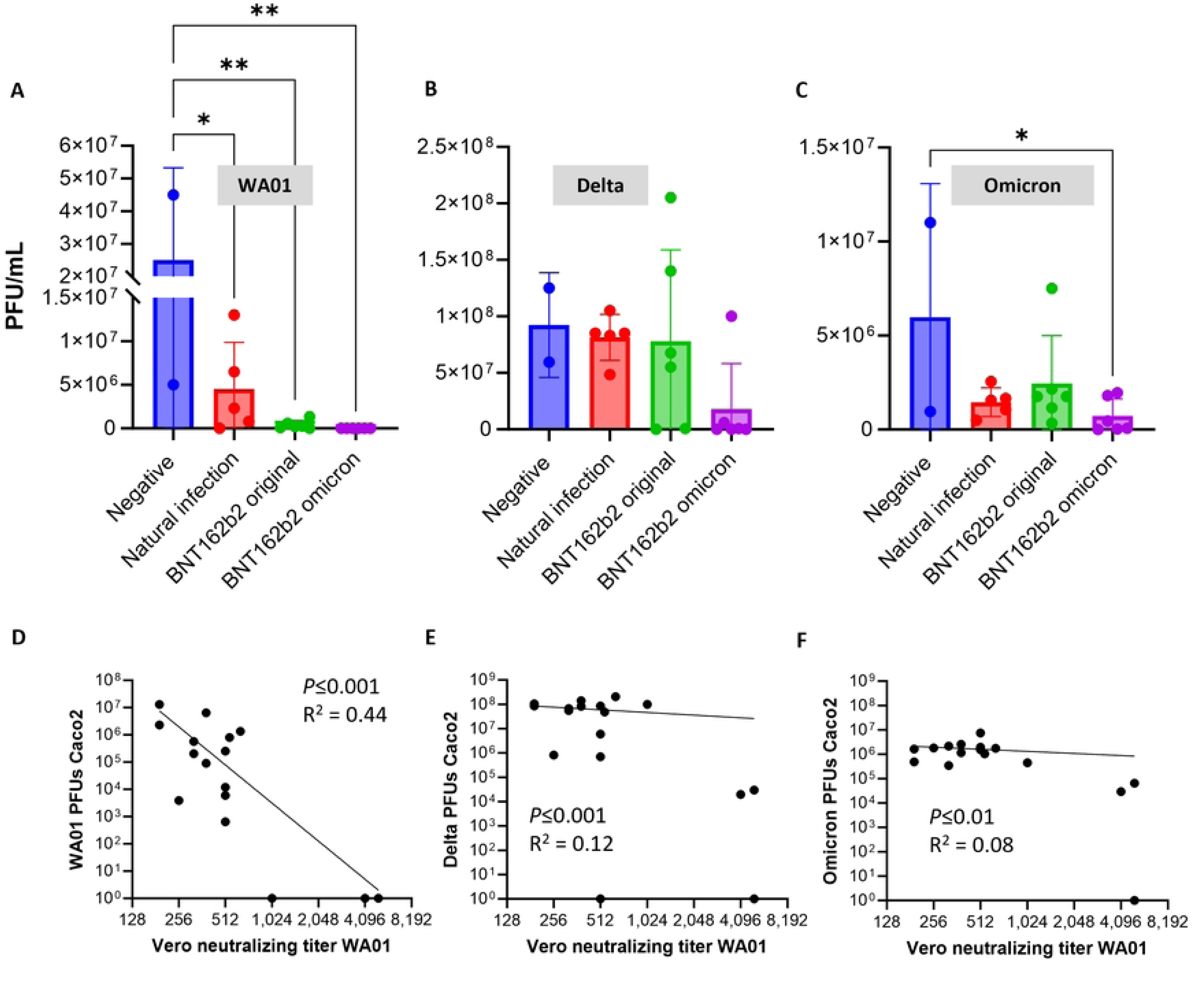
Plaque assay analysis of serum inhibition of SARS-CoV-2 infection in intestinal epithelial cells for strains WA01, Delta and Omicron. SARS-CoV-2 was pretreated for 1 h with negative control sera, sera from individuals who had undergone a natural SARS-CoV-2 infection, or sera from individuals who had received the original or an Omicron-based Pfizer BNT162b2 vaccine. The virus then was inoculated at an MOI=1 onto confluent monolayers of Caco-2 cells. Samples were analyzed after 48 h. (**A-C**) Viral replication of the three SARS-CoV-2 strains was quantified by plaque assay analysis of culture supernatants. Pooled samples from two independent experiments with three technical replicates each are shown. Symbols represent two to six serum samples per group, bars represent mean ± SD. * *P* ≤ 0.05; ** *P* ≤ 0.01; one-way ANOVA with Dunnet’s multiple comparisons test. (**D-E**) Comparison of neutralization titers determined using strain WA01 in VeroE6 cells and ability of each serum to block infection of Caco-2 cells with strains WA01, Delta and Omicron. A simple non-linear correlation analysis was performed, with “zero” values for Caco-2 infection converted to “one”. Each dot represents one serum sample. R^2^ was calculated based on an unconstrained Y intercept.

In contrast, none of the sera significantly inhibited viral replication determined by PFUs with the Delta AY.1 strain of SARS-CoV-2 (**Fig. 3B**). Although a strong trend for decreased PFUs was seen for most sera from donors that had received Omicron-based boosters, one of the samples appeared to enhance viral replication. Similarly, some sera from donors who had received the original BNT162b2 vaccine led to increased replication of the SARS-CoV-2 Delta strain, while other sera prevented viral replication.

Similar to infection with the SARS-CoV-2 Delta strain, immune sera had only limited impact on Caco-2 infection with the Omicron BA.1 strain. Only sera from the Omicron based BNT162b2 vaccine significantly inhibited SARS-CoV-2 infection (**Fig. 3C**). Viral replication was consistently lower in the presence of sera collected after natural infection or vaccination compared to negative sera, but this trend was not significant, likely due to the large variation in viral replication in the presence of negative pre-COVID-19 sera.

Comparison of plaque assay data for all conditions by two-way ANOVA revealed a significant impact of virus strain (*P<*0.001) and a trend for a significant impact of immunization type (*P* = 0.06). Interestingly, the sera that increased replication of Delta (#247 and #298) or Omicron (#263) in Caco-2 cells had moderate to high serum neutralization titers for WA01 in the VeroE6 assay (**Table 1**).

A simple non-linear correlation comparing serum neutralization titers for WA01 in VeroE6 cells to PFUs observed following infection of Caco-2 cell revealed a significant correlation for WA01 infection in Caco-2 cells at *P≤*0.001, but a weak goodness of fit, with an R^2^ of only 0.44 (**Fig. 3D**). For Caco-2 infection with Delta and Omicron, the correlative relationships were even weaker, with R^2^ of 0.12 (*P≤*0.001) and 0.08 (*P≤*0.01), indicating that serum neutralizing activity for WA01 in VeroE6 cells is not a good predictor for protective activity in different cells with a different SARS-CoV-2 strain.

Data from immunofluorescence analysis performed in parallel largely confirmed these observations. 2-way ANOVA analysis comparing the percentage of infected cells revealed a significant effect of virus strain (*P* <0.001) and immunization type *(P* = 0.04). For Caco-2 cells that were exposed to SARS-CoV-2 WA01, the proportion of NP-positive cells was significantly reduced in all samples that contained sera from vaccinated donors (**Fig. 4A-E**). Almost no infected cells were seen when virus was treated with sera from donors who had received an Omicron-based vaccine in 2023 (**Fig. 4D, E**). Consistent with the results from the plaque assays (Fig. 3), none of the sera consistently inhibited Caco-2 infection with Delta, and some sera were associated with an increased number of infected cells (**Fig. 4F**). Similarly, none of the sera significantly reduced infection with Omicron, although a trend was seen for lower infection rates after immunization with the Omicron-based BTN2b2 vaccine (**Fig. 4G**).

**Figure 4:**
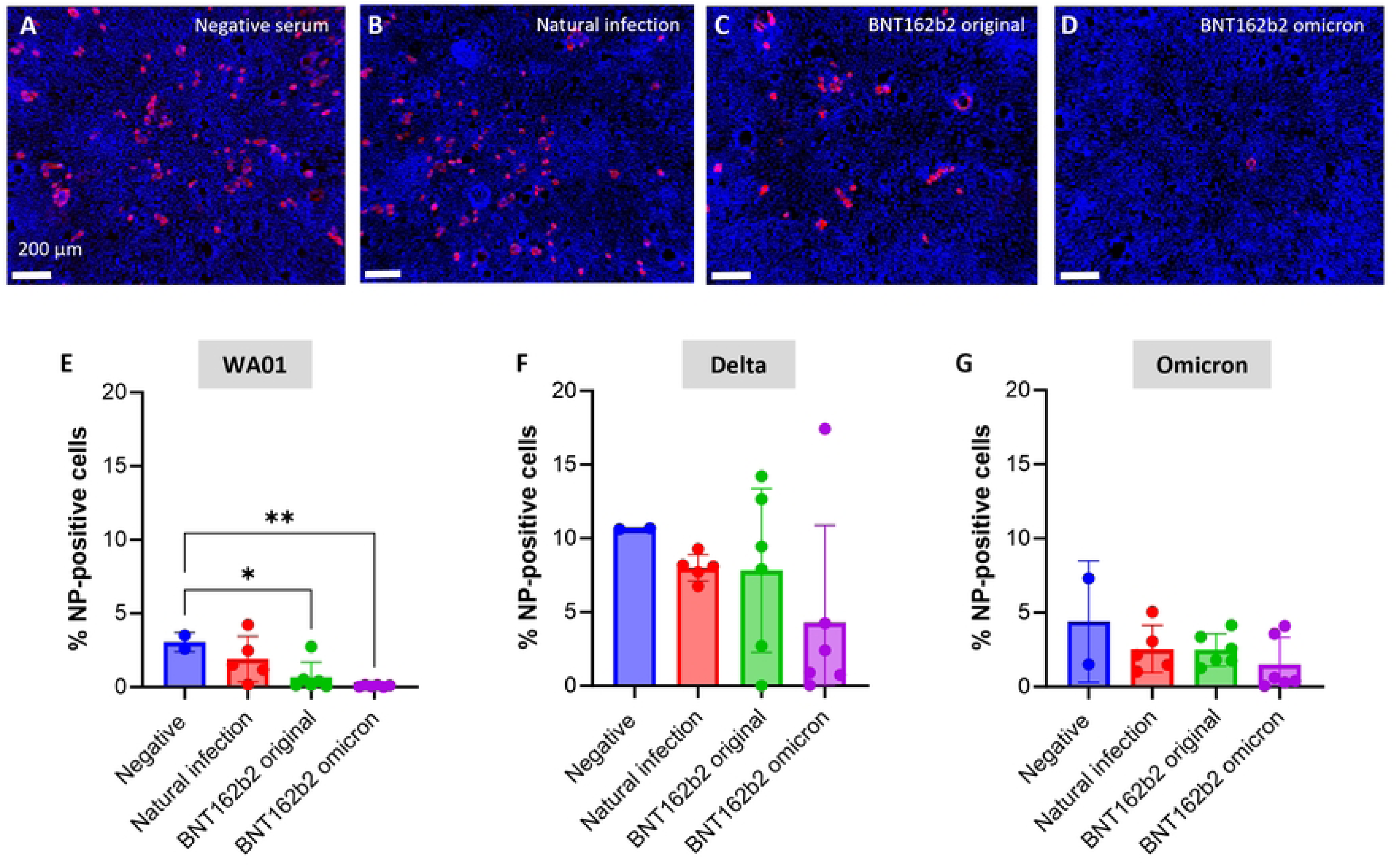
Immunofluorescence analysis of serum inhibition of SARS-CoV-2 infection in intestinal epithelial cells for strains WA01, Delta and Omicron. SARS-CoV-2 was pretreated for 1 h with (**A**) negative control serum, (**B**) serum from an individual who had undergone a natural SARS-CoV-2 infection, or sera from individuals who had received (**C**) the original or (**D**) an Omicron-based Pfizer BNT162b2 vaccine. The virus then was inoculated at an MOI=1 onto confluent monolayers of Caco-2 cells. Samples were analyzed after 48 h. Data represent three technical replicates and two to six biological replicates. Immunofluorescence images of SARS-CoV-2-infected Caco-2 monolayers for viral nuclear protein (Alexa 555, red) were collected by confocal microscopy. DAPI nuclear stain is shown in blue. Bar = 150 µm. (**E, F, G**) Replication of SARS-CoV-2 was measured by immunofluorescence staining of SARS-CoV-2 NP and quantitative image analysis. Pooled samples from two independent experiments are shown. Symbols represent average values of three technical replicate wells for each serum sample that was tested, bars represent mean ± SD. * *P* ≤ 0.05; ** *P* ≤ 0.01; one-way ANOVA with Dunnet’s multiple comparisons test.

## Discussion

In this study, we tested human sera from individuals exposed to SARS-CoV-2 by natural infection or immunization with the Pfizer BNT162b2 COVID-19 vaccines for their ability to block infection of intestinal epithelial cells with different strains of SARS-CoV-2. Our results showed that SARS-CoV-2 immune sera inhibited infection of the intestinal epithelium in two *in vitro* models, but only sera from individuals who had received omicron-based BNT162b2 boosters robustly blocked viral replication in intestinal epithelial cells and were most effective against infection with the original WA01 SARS-CoV-2 strain.

Our experiments using both embryonic stem-cell derived intestinal organoids and Caco-2 cells confirmed previous studies that showed that SARS-CoV-2 replicates robustly in human intestinal epithelial cells [3, 5, 29, 39]. Inducible pluripotent stem cell-derived organoids have emerged as an improved model to study SARS-CoV-2 infection in the human gut, since they replicate the microscopic anatomy of the intestine more faithfully than cell lines or adult stem cell derived organoids [41–44]. Caco-2 cells are a commonly used intestinal epithelial cell line derived from a colon carcinoma that also was shown to support robust replication of SARS-CoV-2, likely due to their high level of ACE2 and TMPRSS2 expression [24, 30]. Interestingly, our immunofluorescence analyses found NP expression in only a small proportion (<10 %) of Caco-2 cells, although viral production determined by plaque assay increased by >2 log fold. Our observations support previous studies that found that enterocytes can produce large quantities of virus [3, 39, 45]. In the human gut, SARS-CoV-2 infected enterocytes also seem to be rare, as shown by us and others [9, 10, 46, 47], but viral detection in the stool is common [46, 48, 49].

These observations suggest that a small number of enterocytes harboring high levels SARS-CoV-2 may still have a major role in the pathogenesis of COVID-19 and the maintenance of a viral reservoir in the context of chronic disease. Notably, there was little evidence for cytopathic effects in the Caco-2 model, again confirming previous reports, although some syncytia formation was observed. Syncytia formation was not strain specific, which differs from previous studies in VeroE6-TMPRSS2 and nasal epithelial cells, where Delta was associated with more significant syncytia formation [50, 51]. Notably, to our knowledge, neither intestinal organoids nor cell lines have to date been used to assess antibody protection from intestinal SARS-CoV-2 infection.

Our results indicate that serum antibodies to SARS-CoV-2 can successfully block infection of human enterocytes, with some exceptions. While these results were expected, we still considered it important to demonstrate epithelial protection from SARS-CoV-2 by antibody neutralization experimentally. Expression of ACE-2 is higher in the intestine than in the lung [52, 53], and it has been suggested that Omicron strains have an increased tropism for the intestine [38]. Therefore, increased amounts of antibody may be needed to block infection. Alternatively, SARS-CoV-2 infection can spread laterally via cell-cell transmission, a mechanism that is likely cell-type specific and that is refractory to antibody neutralization [54].

In addition, intestinal epithelial cells, including Caco-2 cells, express Fc receptors including the neonatal Fc receptor (FcRn) and the polymeric immunoglobulin receptor (pIgR), which bind the constant portions of IgG and IgA antibodies, respectively, and can mediate internalization and transcytosis of immune complexes [55]. It has been suggested that Fc-mediated transport of IgG or IgA immune complexes could lead to antibody-mediated enhancement [55, 56]. The antibody dependent enhancement (ADE) phenomenon has been previously reported for other coronaviruses, such as and Middle East Respiratory Syndrome (MERS)-CoV, SARS-CoV, and feline infectious peritonitis virus infection [55, 57]. Interestingly, Kam et al. showed that spike-specific antisera reduced SARS-CoV pseudoviral entry into cells that were ACE2-positive and FcR-negative, but enhanced viral entry into human B cells that expressed Fc receptors, but lacked ACE2 [58]. FcR-mediated entry also has been demonstrated for SARS-CoV-2 [57]. Our experiments revealed increased replication of Delta with two sera from immunized individuals, and of Omicron with one specific serum, which could potentially represent ADE. However, since we compared sera from vaccinated donors to “negative” sera collected pre-COVID-19, the increased replication of Delta and Omicron in the presence of three specific serum samples could also reflect a lack of variant-specific antibodies coupled with a low concentration of innate antiviral factors in these particular samples. Notably, in most studies, ADE observed *in vitro* does not translate into ADE *in vivo*, and there is currently no solid evidence that ADE contributes to pathogenesis of COVID-19 [57].

Interestingly, we found that sera from individuals who had received Omicron-based BNT162b2 boosters conveyed improved protection from intestinal infection with the original WA01 SARS-CoV-2 strain. This may have been partially due to the somewhat increased antibody levels present in sera from individuals who had received multiple boosters, which was the case for all donors who received the omicron-based BNT162b2. However, the increased antibody titers do not fully explain this phenomenon. Overall, anti-WA01 antibody neutralization titers measured in VeroE6 cells correlated moderately well with protection from Caco-2 infection with the WA01 strain, pointing to contributions of some cell-type specific effects. Conversely, there was very little correlation between VeroE6 neutralization titers and protection of Caco-2 cells from the SARS-CoV-2 strains Delta and Omicron. These observations are consistent with the lack of significant protection of Caco-2 cells from Delta by any of the immune sera, and the protection from Omicron by the Omicron-based vaccine only that we observed and likely represent immune evasion by the new viral variants of concern.

Our experiments with intestinal epithelial cells confirm previous reports that have shown immune escape by SARS-CoV-2 variants of concern, particularly Delta and Omicron. While vaccines were initially highly effective in inducing protection from original SARS-CoV-2 strains [16, 17], the rapid evolution and spread of SARS-CoV-2 led to the emergence of viral variants with mutations in the receptor binding motif of the spike protein [13, 62–64]. For Delta, mutations in the n-terminal domain of spike have been associated with escape from antibody mediated neutralization [59]. Similarly, multiple mutations in the spike sequences of Omicron that are associated with successful evasion of antibody responses that were induced by previous SARS-CoV-2 strains or vaccines have been described [19, 60, 61]. Notably, the sera from infected individuals that we used in this study were all collected early during the pandemic, before the Delta and Omicron variants arose, so that antibodies against these variants we likely not prevalent. Similarly, no Delta-specific sequences were included in any of the BNT162b2 vaccines. Therefore, the results presented in our paper are not unexpected, although additional mechanisms may have contributed to the highly variable responses that we observed.

We are aware that our study had several limitations. First, only a small number of serum samples was included, the epitope binding capacity of the sera was not analyzed in detail, and the samples were collected at different time points after vaccine administration or SARS-CoV-2 infection, and additional natural SARS-CoV-2 infections in vaccinated donors were not systematically tracked, all resulting in highly variable samples. Second, the majority of experiments were performed in the Caco-2 model, since it was more amenable to high throughput screening than the HIOs, although HIOs are likely the more relevant model, since HIOs contain multiple relevant types of primary epithelial cells [27]. A third limitation is that we used serum antibodies to assess mucosal protection in the intestine, since collecting intestinal secretions to assess mucosal antibodies was outside the scope of this study. However, serum antibody levels are commonly used as a correlate of protection for vaccines against intestinal infections [62]. Natural infection with SARS-CoV-2 induces spike-neutralizing antibodies in the serum that are predominantly IgG1, but that also include other IgG subclasses and IgA [63]. At mucosal sites, both IgG and IgA antibodies directed against SARS-CoV-2 spike can be detected following systemic immunization with BNT162b2 [64, 65]. Since we performed functional neutralization assays only, it is unclear which antibody class mediated protection in our study.

Overall, our results indicate that vaccine-induced antibodies reliably prevented infection with the early pandemic strain WA01, but had only limited effects on Delta and Omicron variants. In general, sera from individuals who had received multiple doses of BNT162b2, including booster vaccines based on Omicron sequences, were the most potent at inhibiting replication of any of the viral strains. While protective effects through antibody neutralization were generally similar for VeroE6 and Caco-2 cells when the same viral strain was studied, cell-type specific variations were also observed. In general, our results support previous reports that found that vaccines need to be adapted to specific SARS-CoV-2 strains to achieve significant protection through neutralizing antibodies. Updated vaccines and hence boosters are likely to remain necessary to ensure protection from newly emerging SARS-CoV-2 variants, regardless of the site of infection.

## Data availability

All relevant data are within the manuscript and its Supporting Information files.

## Acknowledgements

We would like to thank all the human volunteers who provided serum samples for this study. Funding from the National Institutes of Health (U01EB029242-02S1 and U01EB029242; to D.B., J.N.W., C.B.C, S.T.W and M.J.), the Montana Agricultural Experiment Station (to D.B. and M.J.), and from the Montana State University Vice President of Research, Economic Development and Graduate Education (M.A.J. and J.H) is gratefully acknowledged.

## Author contributions

Conceptualization: MDC, JH, STW, MAJ, EKL, DB

Formal Analysis: MDC, BS, MAJ, EKL, DB

Funding acquisition: STW, CBC, JNW, MAJ, DB

Investigation: MDC, DS, BS, KB, BJ, AS, JFH, EKL

Methodology: MDC, DS, BS, BJ, SH, JFH, EKL

Project administration: STW, MAJ, EKL, DB

Resources: SH, JH, STW, JRS, MAJ, EKL, DB

Supervision: JH, MAJ, EKL, DB

Visualization: MDC, BS, BJ, MAJ, EKL, DB

Writing – original draft: MDC, DS, KB, EKL, DB

Writing – review & editing: MDC, DS, EKL, DB

